## Supplementary material for "Who would take part in a pandemic preparedness cohort study? The role of vaccine-related affective polarisation: cross-sectional survey": S1 Figure

### S1 figure: Distribution of raw data from variables which were dichotomised

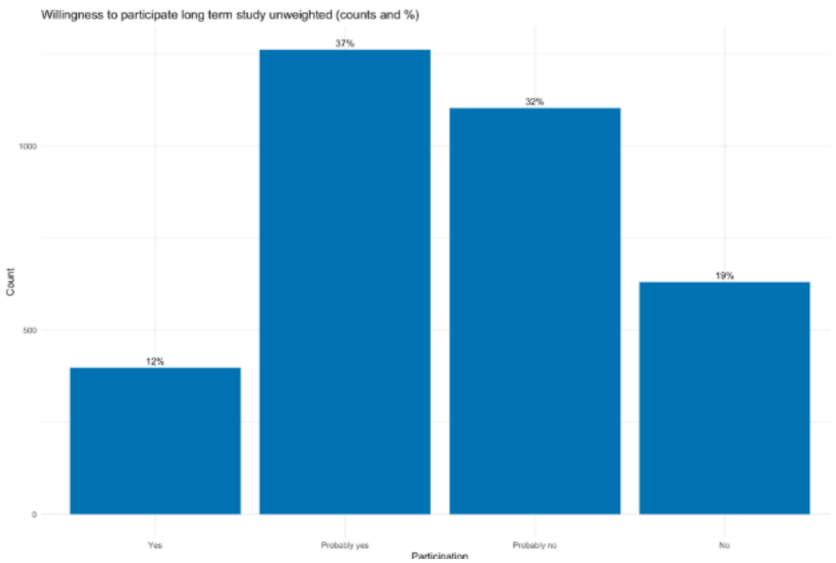


**1a:** Responses to, “In principle, would you be willing to participate in a long-term study?”

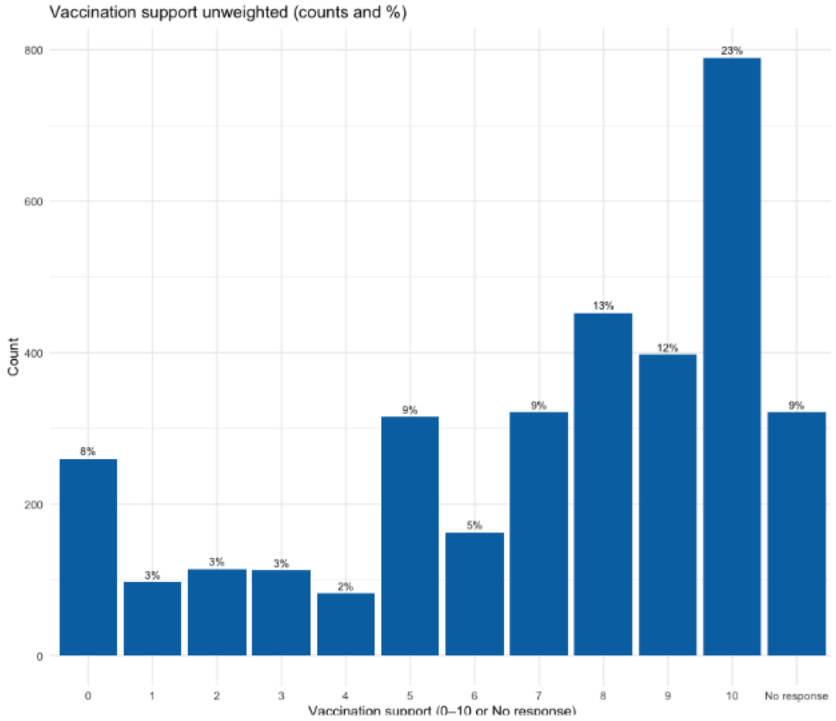


**1b:** Responses to “What do you think about vaccinations against COVID-19?”

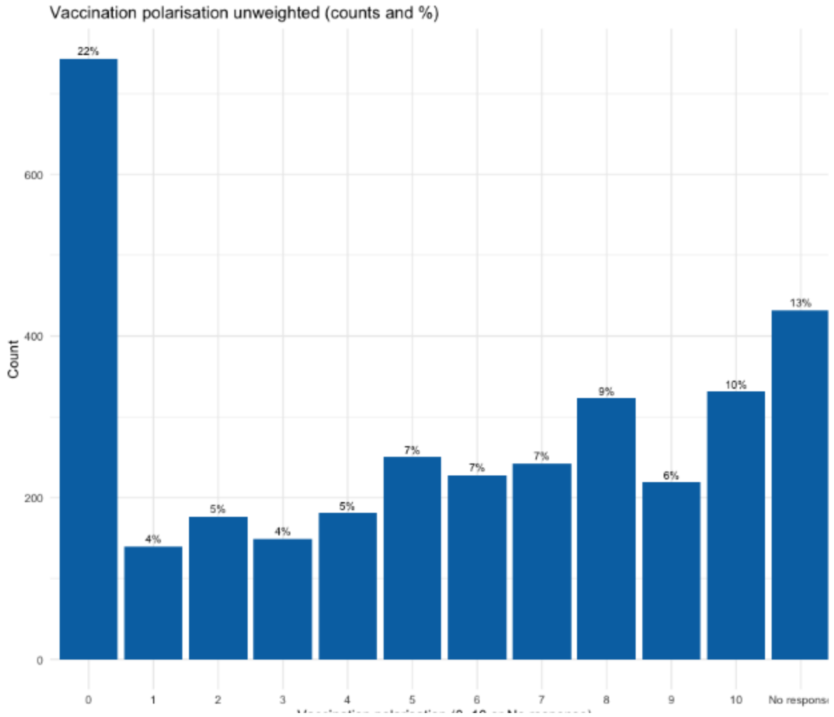


**1c:** Affective polarisation was calculated as the absolute difference between participants' feelings toward people who get vaccinated and those who do not, each rated on a scale from -5 (very negative) to +5 (very positive). The resulting score ranges from 0 to 10, where 0 indicates no polarisation and 10 indicates strong affective polarisation.
