## Supplementary material for "Who would take part in a pandemic preparedness cohort study? The role of vaccine-related affective polarisation: cross-sectional survey": S2 Figure

### S2 figure: Study participants flow chart

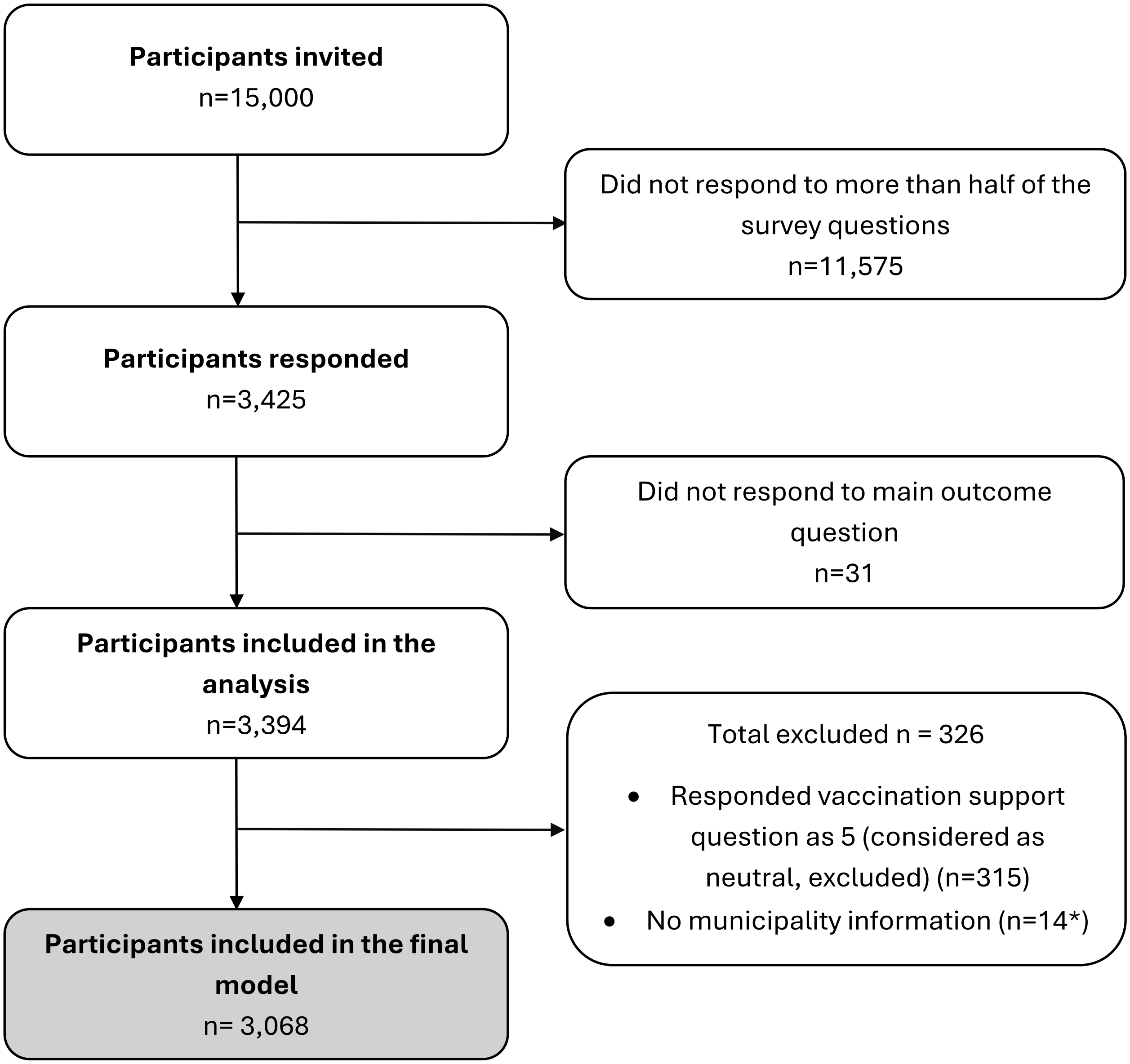


*3 missing values in “no municipality information” overlaps with responded vaccination as 5.
