## Supplementary material for "Who would take part in a pandemic preparedness cohort study? The role of vaccine-related affective polarisation: cross-sectional survey": S2 Table

### S2 table: Descriptive table for variables included in the analysis

| Variable Name | Survey Question Asked | Survey answers | Analysis |
|---|---|---|---|
| Households size | Number of people in your household (including yourself) | 1 to 20 | Categorical variable<br>1<br>2<br>3<br>4<br>5 or more |
| Education level | What is your highest level of education? | 1) Primary school<br>2) Secondary school 3) Grammar school 4) Vocational training/Federal Vocational Baccalaureate<br>5) University degree: Bachelor<br>6) University degree: Master/Diploma<br>7) University degree: Doctorate/PhD<br>8) Other | Categorical variable<br>Compulsory or less (1 and 2)<br>Upper secondary (3 and 4)<br>Tertiary education (5, 6 and 7)<br>Other (8)<br>No response |
| Age in years | Age person | 18 to 120 years | Continuous variable |
| House location | Where is your flat/house located? | 1) City centre<br>2) Suburb<br>3) Industrial area<br>4) Rural area<br>5) Adjacent to farmland<br>6) Other | Categorical variable<br>City centre (1)<br>Suburb (2)<br>Rural area (4 and 5)<br>Other (3 and 6) |
| Income, Swiss Francs | What is your average net household income per month? | 1) <3,000<br>2 3,000-4,500<br>3) 4,500-6,000<br>4) 6,000-9,000<br>5) 9,000-11,000<br>6) 11,000 or more<br>7) Prefer not to say<br>8) Don't know | Categorical variable<br><4,500 (1 and 2)<br>4,500-9,000 (3 and 4)<br>>9,000 (5 and 6)<br>Other (7 and 8)<br>No response |
| Current work situation | How would you best describe your current labour situation? | 1) Working full-time (80%+)<br>2) Full-time housewife/-husband (80% or more)<br>3) Working part-time (<80%)<br>4) Casual worker<br>5) Unemployed<br>6) Never worked<br>7) In education<br>8) Long-term absence, i.e. more than 3 months (unpaid leave, maternity leave etc.)<br>9) Not working due to health reasons (incl. Disability benefits)<br>10) Pensioner<br>11) Pensioner but working (incl. in an honorary capacity)<br>12) Volunteer | Categorical variable<br>Full time employee (1)<br>Part time employee (3,4 and 11)<br>Not employed (2, 5, 6,8,9,10 and 12)<br>In education (7)<br>Other (13)<br>No response |

| | | | |
|---|---|---|---|
| | | 13) Other | |
| Gender | I am ... | 1) Male<br>2) Female<br>3) Non-binary<br>4) Prefer not to say<br>5) Don't know | Categorical variable<br>Female (2)<br>Male (1)<br>Other (3,4 and 5)<br>No response |
| Language | Languages | 1) Deutsch<br>2) Français<br>3) Italiano<br>4) English | Categorical variable<br>German<br>French<br>Italian<br>English |
| Nationality | What's your nationality? | 1) Swiss<br>2) Foreign nationality<br>3) More than one nationality<br>4) Prefer not to say<br>5) Don't know | Categorical variable<br>Swiss<br>Foreign<br>No response |
| Feelings about COVID-19 vaccination | What do you think about vaccinations against COVID-19? | 0 to 10<br>(0, I completely reject them to 10, I completely support them)<br>Slider used to set response | Categorical variable<br>Vaccination supporter (>5)<br>Vaccination opposer (<5)<br>(5 neither supporter or opposer) |
| Feeling thermometer questions | Some people do not want to get vaccinated while others get vaccinated against COVID-19. Now, please think about these two groups. How do you feel about people belonging to these two groups? Please describe your feelings using a thermometer ranging from-5 to +5. -5 means that you are very cold and negative towards the group. +5 means that you are very sympathetic and positive towards the group. | | |
| Feelings about people who get vaccinated | People who get vaccinated | -5 to +5<br>-5 very cold and negative<br>+5 very warm and positive<br>Change the slider above to set a response. | Not directly part of the model<br>“Vaccination related affective polarisation” variable calculated with this variable. |
| Feelings about people who do not get vaccinated | People who do not get vaccinated | -5 to +5<br>-5 very cold and negative<br>+5 very warm and positive<br>Slider used to set response | Not directly part of the model<br>“Vaccination related affective polarisation” variable calculated with this variable. |
| Vaccination related affective polarisation (Calculated) | Not a survey question calculated using “Feelings about people who get vaccinated” and “Feelings about people who do not get vaccinated” | Not a survey question calculated using “Feelings about people who get vaccinated” and “Feelings about people who do not get vaccinated” | Binary variable<br>Polarised<br>Not polarised |
| Willingness to participate long term study | In principle, would you be willing to participate in a long-term study | 1) Not at all<br>2) Yes, definitely<br>3) Rather yes<br>4) Rather no | Binary variable (Outcome)<br>Yes (2 and 3)<br>No (1 and 4) |

| What would be your motivation(s) to participate in a long-term study and to donate time, information and potentially biological samples to it? (responders can choose multiple answers from down below) |
|---|
| In this way I can contribute to the health of fellow human beings |
| I can contribute to better preparation for the next pandemic |
| I am interested in research and health |
| I get a free health check |
| I receive the study results as feedback |
| It makes me proud to be a participant in an important cantonal study |
| Other reasons (free text option) |

| For what reasons would you refuse participation in a long-term study? | Assigned categories in figure 2 |
|---|---|
| I am not interested in it | Not interested |
| I don't think much of health research | Not interested |
| I don't want to share my health data | Privacy concerns |
| I am worried that my data might not be sufficiently well protected | Privacy concerns |
| I am worried that my data may be abused (e.g. by health insurers, employers or similar) | Privacy concerns |
| I don't want to donate blood or other biological samples | Mistrust |
| I am worried that my voluntary contribution may serve the private interests of the pharmaceutical industry | Mistrust |
| I don't believe that such a study will improve the health of the population | Mistrust |
| I have no time | No time |
| I only have time in the evening and on weekends to participate in a study | No time |
| I don't want to visit a study centre | No benefit |
| I don't personally benefit from the results of such a study | No benefit |
| Prefer not to say | Other reasons / no response |
| Other reasons | Other reasons / no response |
| Don't know | Other reasons / no response |
