## Supplementary material for "Who would take part in a pandemic preparedness cohort study? The role of vaccine-related affective polarisation: cross-sectional survey": S3 Table

### S3 table: Weighting levels and represented households

We applied weighting to our dataset at two different levels. At the first level, we weighted all invited participants to conduct a descriptive analysis comparing responders and non-responders. At the second level, we weighted only the participants who responded in order to perform our primary logistic regression analysis.

| Level 1: Invited participants | | | Level 2: Responded participants | | |
|---|---|---|---|---|---|
| Household size | Study sample (invited) | Households represented | Household size | Study sample (respondents) | Households represented |
| 1 | 3,000 | 185,820 | 1 | 639 | 185,820 |
| 2 | 3,000 | 170,629 | 2 | 877 | 170,629 |
| 3 | 3,000 | 54,814 | 3 | 718 | 54,814 |
| 4 | 3,000 | 53,981 | 4 | 809 | 53,981 |
| 5+ | 3,000 | 24,729 | 5+ | 351 | 24,729 |
