## Supplementary material for "Who would take part in a pandemic preparedness cohort study? The role of vaccine-related affective polarisation: cross-sectional survey": S4 Table

### S4 table: Responders of the survey by household size and language

| | Response to survey | | | | | | | | |
|---|---|---|---|---|---|---|---|---|---|
| | Yes | | | No | | | Total | | |
| | N[1] | Col%[2] | Row%[3] | N[1] | Col%[2] | Row%[3] | N[1] | Col%[2] | Row%[3] |
| **Household size** | | | | | | | | | |
| 1 | 639 | 18.8 | 21.3 | 2,361 | 20.3 | 78.7 | 3,000 | 20 | 100 |
| 2 | 877 | 25.8 | 29.2 | 2,123 | 18.3 | 70.8 | 3,000 | 20 | 100 |
| 3 | 718 | 21.2 | 23.9 | 2,282 | 19.7 | 76.1 | 3,000 | 20 | 100 |
| 4 | 809 | 23.8 | 27.0 | 2,191 | 18.9 | 73.0 | 3,000 | 20 | 100 |
| 5+ | 351 | 10.3 | 11.7 | 2,649 | 22.8 | 88.3 | 3,000 | 20 | 100 |
| **Language** | | | | | | | | | |
| German | 3,074 | 90.6 | 22.8 | 10,400 | 89.6 | 77.2 | 13,474 | 89.8 | 100 |
| French | 312 | 9.2 | 21.2 | 1,161 | 10.0 | 78.8 | 1,473 | 9.8 | 100 |
| Italian | 2 | 0.06 | 18.2 | 9 | 0.08 | 81.8 | 11 | 0.07 | 100 |
| English | 6 | 0.17 | 14.6 | 35 | 0.30 | 85.4 | 41 | 0.23 | 100 |

[1] Number of observations
[2] Column percentage
[3] Row percentage
