## Supplementary material for "Who would take part in a pandemic preparedness cohort study? The role of vaccine-related affective polarisation: cross-sectional survey": S5 Table

### S5 table: Willingness to participate in a long-term cohort study, unweighted denominators and proportions

| | | Denominator | Willingness to participate long term study |
|---|---|---|---|
| **Responders** | | 3,394 | 1,660 |
| **Age, years (SD; range)** | | 3,394 | 48.6 (16.5; 18-94) |
| **Age group, years** | 18-29 | 404 (11.9%) | 223 (55.2%) |
| | 30-64 | 2,261 (66.6%) | 1,132 (50.0%) |
| | 64+ | 729 (21.4%) | 305 (41.8%) |
| **Gender** | Female | 1,787 (52.7%) | 899 (50.3%) |
| | Male | 1,580 (46.6%) | 750 (47.5%) |
| | Other | 13 (0.4%) | 7 (53.8%) |
| | No response | 14 (0.4%) | 4 (28.6%) |
| **Education level** | Compulsory education or less | 488 (14.4%) | 153 (31.4%) |
| | Upper secondary | 1,522 (44.8%) | 668 (43.9%) |
| | Tertiary | 1,300 (38.3%) | 808 (62.2%) |
| | Other | 26 (0.8%) | 10 (38.5%) |
| | No response | 58 (1.7%) | 21 (36.2%) |
| **Current work situation** | Full-time employee | 1,473 (43.4%) | 770 (52.3%) |
| | Part-time employee | 815 (24%) | 439 (53.9%) |
| | Not employed | 830 (24.5%) | 336 (40.5%) |
| | In education | 127 (3.7%) | 73 (57.5%) |
| | Other | 56 (1.6%) | 19 (33.9%) |
| | No response | 93 (2.7%) | 23 (24.7%) |
| **Income, Swiss Francs** | <4,500 | 518 (15.3%) | 213 (41.1%) |
| | 4,500 – 9,000 | 1,349 (39.8%) | 666 (49.4%) |
| | >9,000 | 911 (26.8%) | 583 (64.0%) |
| | Other | 480 (14.1%) | 161 (33.5%) |
| | No response | 136 (4.0%) | 37 (27.2%) |
| **Household size** | 1 | 639 (18.8%) | 336 (52.6%) |
| | 2 | 877 (25.8%) | 423 (48.2%) |
| | 3 | 718 (21.2%) | 367 (51.1%) |
| | 4 | 809 (23.8%) | 392 (48.5%) |
| | 5+ | 351 (10.3%) | 142 (40.5%) |
| **Household location[1]** | Urban | 1,864 (54.9%) | 970 (52.0%) |
| | Intermediate | 861 (25.4%) | 405 (47.0%) |
| | Rural | 655 (19.2%) | 279 (42.6%) |
| **Nationality** | Swiss | 2,716 (80.0%) | 1,377 (50.7%) |

| | | | |
|---|---|---|---|
| | Foreign | 637 (18.8%) | 273 (42.9%) |
| | No response | 41 (1.2%) | 10 (24.4%) |
| **Language** | German | 2,922 (86.1%) | 1,435 (49.1%) |
| | French | 330 (9.7%) | 154 (46.7%) |
| | Italian | 49 (1.4%) | 22 (44.9%) |
| | English | 93 (2.7%) | 49 (52.7%) |
| **Vaccination support status**[2] | For vaccination | 2,106 (62.0%) | 1,220 (57.9%) |
| | Against vaccination | 655 (19.3%) | 210 (32.1%) |
| | No response | 321 (9.5%) | 102 (31.8%) |
| **Vaccination related affective polarisation** | Polarised | 1,376 (40.5%) | 607 (44.1%) |
| | Not polarised | 1,586 (46.7%) | 893 (56.3%) |
| | No response | 432 (12.7%) | 160 (37.0%) |
| **Willingness to participate with children** | | | |
| | | **Number with children** | **Willingness to participate** |
| | | | Yes |
| **On behalf of their children** | - | 1,083 | 450 (41.6%) |
| **Willingness to participate with pets** | | | |
| | | **Number with pets** | **Willingness to participate** |
| | | | Yes |
| **On behalf of their pets** | Any pet | 1,066 | 594 (55.7%) |
| **Type of Pet**[3] | Dogs | 336 (31.5%) | 158 (47%) |
| | Cats | 710 (66.6%) | 312 (43.9%) |
| | Rabbits | 83 (7.8%) | 34 (41%) |
| | Rodents | 71 (6.6%) | 38 (53.5%) |
| | Others | 124 (11.6%) | 56 (45.2%) |

[1] The sum of percentages for the household location variable does not equal 100% due to missing data that were not classified (n=14) by the Cantonal Administration and Information Office.

[2] The sum of percentages for the opinions on vaccination does not total 100% because responses marked as “5” (n=312) - representing a neutral position at the midpoint of the scale - were excluded from the analysis.

[3] The sum of the percentages exceeds 100% because some households reported owning multiple types of pets.
