## Supplementary material for "Who would take part in a pandemic preparedness cohort study? The role of vaccine-related affective polarisation: cross-sectional survey": S6 Table

### S6 table: Sensitivity analysis

| | Multivariable (final) model | | Sensitivity analysis model 1 | | Sensitivity analysis model 2 | | Sensitivity analysis model 3 | |
|---|---|---|---|---|---|---|---|---|
| **Characteristic** | **OR[1]** | **95% CI[1]** | **OR[1]** | **95% CI[1]** | **OR[1]** | **95% CI[1]** | **OR[1]** | **95% CI[1]** |
| **Age, per year** | 0.99 | 0.98 - 0.99 | 0.99 | 0.98 - 0.99 | 0.99 | 0.98 - 0.99 | 0.99 | 0.98 - 0.99 |
| **Education level** | | | | | | | | |
| Compulsory education or less | 1 | 1 | 1 | 1 | 1 | 1 | 1 | 1 |
| Upper secondary education | 1.59 | 1.19 - 2.12 | 1.53 | 1.16 - 2.02 | 1.54 | 1.17 - 2.03 | 1.59 | 1.19 - 2.12 |
| Tertiary education | 2.48 | 1.81 - 3.39 | 2.41 | 1.79 - 3.24 | 2.39 | 1.77 - 3.23 | 2.46 | 1.80 - 3.37 |
| Other | 1.57 | 0.49 - 5.05 | 1.36 | 0.49 - 3.77 | 1.54 | 0.55 - 4.35 | 1.56 | 0.48 - 5.07 |
| No response | 1.61 | 0.75 - 3.43 | 1.41 | 0.68 - 2.91 | 1.43 | 0.71 - 2.89 | 1.62 | 0.76 - 3.47 |
| **Income, Swiss Francs** | | | | | | | | |
| <4'500 | 1 | 1 | 1 | 1 | 1 | 1 | 1 | 1 |
| 4'500 - 9'000 | 1.21 | 0.93 - 1.59 | 1.18 | 0.92 - 1.53 | 1.18 | 0.91 - 1.52 | 1.22 | 0.94 - 1.60 |
| >9'000 | 1.92 | 1.39 - 2.65 | 1.74 | 1.28 - 2.36 | 1.75 | 1.29 - 2.39 | 1.96 | 1.41 - 2.70 |
| Other | 0.62 | 0.44 - 0.87 | 0.60 | 0.43 - 0.83 | 0.61 | 0.44 - 0.85 | 0.62 | 0.44 - 0.87 |
| No response | 0.46 | 0.26 - 0.83 | 0.56 | 0.33 - 0.94 | 0.57 | 0.33 - 0.98 | 0.46 | 0.26 - 0.82 |
| **Number of household members** | | | | | | | | |
| 1 | 1 | 1 | 1 | 1 | 1 | 1 | 1 | 1 |
| 2 | 0.74 | 0.59 - 0.95 | 0.75 | 0.59 - 0.94 | 0.75 | 0.60 - 0.95 | 0.74 | 0.58 - 0.95 |
| 3 | 0.65 | 0.50 - 0.85 | 0.69 | 0.53 - 0.89 | 0.67 | 0.52 - 0.87 | 0.65 | 0.49 - 0.85 |
| 4 | 0.50 | 0.38 - 0.67 | 0.55 | 0.42 - 0.71 | 0.54 | 0.42 - 0.71 | 0.51 | 0.38 - 0.67 |
| 5≤ | 0.67 | 0.44 - 1.01 | 0.66 | 0.45 - 0.98 | 0.66 | 0.45 - 0.97 | 0.66 | 0.43 - 0.99 |
| **Opinion about vaccination** | | | | | | | | |
| Support vaccination | 1 | 1 | 1 | 1 | 1 | 1 | 1 | 1 |
| Oppose vaccination | 0.53 | 0.39 - 0.72 | 0.61 | 0.45 - 0.81 | 0.55 | 0.42 - 0.72 | 0.52 | 0.38 - 0.73 |
| No response | 0.34 | 0.07 - 1.69 | 0.39 | 0.08 - 1.83 | 0.34 | 0.07 - 1.60 | 0.55 | 0.11 - 2.82 |
| **Vaccination polarisation** | | | | | | | | |
| Not polarised | 1 | 1 | 1 | 1 | 1 | 1 | 1 | 1 |
| Polarised | 1.51 | 1.20 - 1.89 | 1.69 | 1.38 - 2.07 | 1.52 | 1.22 - 1.91 | 1.48* | 1.15 - 1.92 |
| No response | 0.88 | 0.47 - 1.67 | 1.11 | 0.63 - 1.95 | 0.89 | 0.47 - 1.68 | 0.89 | 0.47 - 1.68 |

| | | | | | | | | |
|---|---|---|---|---|---|---|---|---|
| **Polarisation x Oppose to vaccination[3]** | 1 | 1 | 1 | 1 | 1 | 1 | 1 | 1 |
| Opposed to vaccination * Polarised | 0.33 | 0.19 - 0.57 | 0.29 | 0.17 - 0.50 | 0.41 | 0.25 - 0.66 | 0.28 | 0.14 - 0.57 |

[1] OR, odds ratio, CI, confidence interval

Multivariable (final) model is the main model that we presented in our manuscript, includes following variables: Age, gender, education level, current work situation, income, household size, household location, language, nationality, opposition to vaccination, polarisation:vaccination opposer(interaction term) and willingness to participate (Outcome). And vaccination support == 5 considered as neutral and excluded

Sensitivity analysis model 1: Includes same variables as the final model however vaccination support == 5 coded as vaccine support instead of being excluded

Sensitivity analysis model 2: Includes same variables as the final model however vaccination support == 5 coded as vaccine oppose instead of being excluded

Sensitivity analysis model 3: Includes same variables as the final model however affective polarisation coded as (≤3 = not polarised, 4 to 6 polarised, 7 and more heavily polarised)

*OR of heavily polarised group (7 and more)
