## Supplementary material for "Who would take part in a pandemic preparedness cohort study? The role of vaccine-related affective polarisation: cross-sectional survey": S7 Table

### S7 table: Reasons to participate or decline long-term study, unweighted

| | |
|---|---|
| **Reasons to participate long-term study** | |
| In this way I can contribute to the health of fellow human beings | 1,559 (56%) |
| I can contribute to better preparation for the next pandemic | 1,134 (41%) |
| I am interested in research and health | 1,031 (37%) |
| I get a free health check | 713 (26%) |
| I receive the study results as feedback | 682 (25%) |
| It makes me proud to be a participant in an important cantonal study | 209 (8%) |
| Other reasons | 115 (4%) |
| **Reasons to decline participating long-term study** | |
| I am not interested in it | 266 (42%) |
| I have no time | 209 (33%) |
| I don't want to share my health data | 133 (21%) |
| I don't want to donate blood (or other biological samples) | 98 (16%) |
| I don't want to visit a study centre | 81 (13%) |
| Prefer not to say | 78 (12%) |
| I am worried that my voluntary contribution may serve the private interests of the pharmaceutical industry | 77 (12%) |
| I don't believe that such a study will improve the health of the population | 76 (12%) |
| I am worried that my data might not be sufficiently well protected | 74 (12%) |
| I am worried that my data may be abused (e.g. by health insurers, employers or similar) | 64 (10%) |
| Other reasons | 45 (7%) |
| I don't personally benefit from the results of such a study | 27 (4%) |
| Don't know | 27 (4%) |
| I don't think much of health research | 15 (2%) |
| I only have time in the evening and on weekends to participate in a study | 12 (2%) |

People who answered to reasons to participate question are those who answered “yes”, “rather yes” and “rather no” to willingness to participate long-term study question.
People who answered to reasons to decline participation question are those who answered “no” to willing to participate long-term study question.
The percentage totals exceed 100% because respondents could select multiple answers for these questions.
